# Safety and Efficacy of Antiviral Drugs for the Treatment of Patients with SARS-CoV-2 Infection: A Systematic Review and Meta-analyses

**DOI:** 10.1101/2020.09.03.20187526

**Authors:** Zuleika Aponte Torres, Sandra Lopez Leon, Thirumugam Muthuvel, Subha Manivannan, Krutika Srivastava, Marco Pavesi

## Abstract

**Objective:** To systematically review the safety and efficacy outcomes of using antivirals for the treatment of COVID-19.

**Methods:** Five databases were screened from inception to 27-Aug-2020. The effects of specific drug interventions on safety and efficacy were assessed in COVID-19 patients. Risk Ratios (RRs) with corresponding 95% confidence intervals (CIs) were pooled using random-effects models.

**Results:** A total of 10 studies were identified which fulfill the inclusion criteria. Patients taking antivirals had 26% less risk of having a severe adverse event (SAE) compared to controls (RR, 0.74, CI:0.62 to 0.89, P=0.002). Clinical improvement at day 14 was observed among the cases treated with antivirals compared to the control group (RR 1.24, CI: 1.00 to 1.53 p=0.05).

**Conclusion:** There is evidence that Remdesivir and LPV/r reduces the hospital length of stay and that patients to which antivirals were administered had less SAE and improvement when compared to patients not prescribed with antivirals. Due to a lack of power and the quality of the studies, it was not possible to determine which antivirals have a greater risk-benefit balance, and therefore the optimal approach to antiviral treatment is still uncertain.

## INTRODUCTION

The novel coronavirus disease (COVID-19) is caused by the SARS-CoV-2 viral pathogen. One of the characteristics of COVID-19 is that it is highly contagious, as of August 27 it has caused more than 24.2 million cases and over 826,743 deaths in 188 countries worldwide. The number of cases of COVID-19 continues to increase but there is no approved vaccine or medication that can be used for treatment.

The repurposed antivirals that are being used as investigational drugs to treat COVID-19 are Favipiravir, Umifenoviridol, a combination of Lopinavir-ritonavir (LPV/r), and Remdesivir (Sanders et al., 2020). These antivirals have been shown to have activity against SARS-CoV-2 in vitro (McKee et al., 2020). Favipiravir and umifenovir are available in Russia and some Asian countries for the treatment of influenza (Wu et al., 2020). Lopinavir-ritonavir is a combined protease inhibitor, which has primarily been used for HIV infection. To date, Remdesivir is the only antiviral to receive emergency approval from the Food Drug Administration (FDA) (Gilead Sciences, n.d., p. 19) in the United States and exception approval from the Japanese Ministry of Health, Labour and Welfare (MHLW) (Gilead Sciences, n.d., p. 19) to treat adults or children who are hospitalized with suspected or confirmed Covid-19 and whose condition is “severe”.

The objective of this systematic literature review (SLR) and the meta-analyses is to assess the safety and efficacy of the antiviral drugs that have been proposed to treat COVID-19. The multiple uncertainties about COVID-19 and a large amount of ongoing research make it necessary to provide the scientific community with high-quality, timely, and living systematic reviews of the relevant evidence.

## METHODS

The protocol for this systematic review and the meta-analyses was based on the PRISMA statement. This protocol has been registered in the PROSPERO database with registration number CRD42020184247. We included all completed and published clinical studies that evaluated the role of antiviral drugs on SARS-CoV-2 or COVID-19. The literature search was conducted in PubMed, EMBASE, Medrxiv, Cochrane Central Register of Controlled Trials (CENTRAL), and LitCOVID databases from inception to 27-Aug-2020 to find articles providing information on the efficacy and safety of antiviral drugs in patients with SARS-CoV-2. No language restrictions were imposed and the search was expanded using a snowballing method applied to the references of retrieved papers.

The exclusion criteria consisted of studies that did not report clinical data. Two authors (ZAT and TM) independently screened the titles and abstracts and the full-texts of potentially relevant articles, using Rayyan software (Ouzzani et al., 2016). Three authors extracted data using a custom spreadsheet to record the study characteristics and study outcomes (mortality, total adverse events (AE), serious adverse events (SAE), and time to a negative PCR test).

Our primary analysis was focused on the safety and efficacy outcomes of the different antiviral therapies proposed for the treatment of patients with COVID-19.

## Risk of bias assessment and quality of the evidence assessment

For Randomized Clinical Trials (RCTs) we used the Cochrane Risk-of-Bias (RoB) assessment tool consisting of seven domains: random sequence generation, allocation concealment, blinding of participants and personnel, blinding of outcome assessment, incomplete outcome data, selective outcome reporting, and other bias (Higgins et al., 2011). For cohort studies, we used the Newcastle-Ottawa Scale (NOS) consisting of three domains (selection of exposure, comparability, and assessment of outcome) (Wells GA,Shea B,O’Connell D, et a1., n.d.). Two reviewers assessed the risk of bias of the articles independently with the good inter-rater agreement (κ =0.75; p<0.001). If the two reviewers did not reach consensus, a third reviewer made the final decision.

## Statistical analysis

Meta-analyses for all outcomes were first performed by pooling the results of all antivirals. Meta-analyses for specific drugs were performed if two or more studies were identified. To assess safety, AE rates were pooled and a list of adverse events for each drug was created (Supplementary Table 2). To assess efficacy, the following outcomes were included in separate meta-analyses: clinical improvement rates at day 7, 14, & 28, chest CT-improvement and 28-days mortality wherever feasible. All meta-analyses were carried out using the RevMan software. All of the outcomes included in the meta-analysis were binary and were summarized using risk ratio (RR) and 95% CI using a random-effects model. Both peer-reviewed as well as non-peer-reviewed studies were pooled in the meta-analyses. I2 testing was used to quantify heterogeneity between studies, values > 50% represented moderate-to-high heterogeneity. Statistical analysis with a p-value < 0.05 was considered statistically significant.

## RESULTS

The electronic searches identified 379 records. Following the screening of titles and abstracts and removing duplicates, we evaluated 20 relevant full text articles that fulfilled the inclusion criteria. After full-text review, an additional 6 articles were excluded, 4 because the studies were ongoing and 2 because they did not include the outcomes of interest. Finally, we found 14 relevant articles suitable for final inclusion, 10 of the studies were included in the meta-analyses (4 RCTs and 6 observational studies). The remaining studies were not included in the analysis because they didn’t fulfil the inclusion criteria.

## Risk of bias assessment

We used the Cochrane risk-of-bias tool for the RCTs evaluation of the risk of bias. The risk of bias in the included RCTs was high, as they did not perform optimal allocation concealment and blindness for patients and clinicians. Wang et al was the study with the highest quality given that it included randomization and allocation concealment; however, it presented insufficient statistical power to detect real differences in the outcomes (Wang et al., 2020). All of the cohort studies had a high risk of bias, the main reasons were the lack of controlling for important factors that would influence the primary study results, lack of long enough follow-up for outcomes to occur, and inadequate outcome ascertainment. Another source of bias was the small sample size in the studies.

## Study characteristics and participant characteristics of the studies included in the meta-analysis Randomized controlled trials (RCTs)

A total of 6 RCT studies were included in the meta-analysis, with a total of 1,784 (951 cases and 833 controls) patients evaluated. All of the included antiviral studies included either LPV/r, LPV/r + Umifenoviridol, Remdesivir, or Favipiravir as the intervention or comparator, the duration of the therapy ranged from 5 days (Wang et al., 2020) to 21 days (Li et al., 2020). LPV/r and Ritonavir doses were similar, the doses used in the study by Cao et al and in the study by Li et al., patients in the Li et al study were given a dose of LPV/r of 200mg/50mg (Li et al., 2020) compared to 400mg/100mg for patients in the Cao et al study (Cao et al., 2020). The mean age of the cases ranged from 58 to 66 years of age compared to controls whose mean age ranged from 58 to 64 years of age (Cao et al., 2020; Wang et al., 2020).

## Overall Outcomes

### Adverse events (AE)

There were five studies that reported AE (Beigel et al., 2020; Cao et al., 2020; Chen et al., 2020; Li et al., 2020; Wang et al., 2020). In all trials (Umifenoviridol, Favipiravir, LPV, Remdesivir) there were gastrointestinal and liver AE reported, except for one trial (Beigel et al., 2020). Umifenoviridol and Favipiravir had no SAE. Serious adverse events (SAE) present in LPV/r and Remdesivir were acute kidney injury, acute respiratory distress syndrome, respiratory failure, hemorrhage of the lower digestive tract, and septic shock. Figure 1 shows the most common AE.

**Figure 1:**
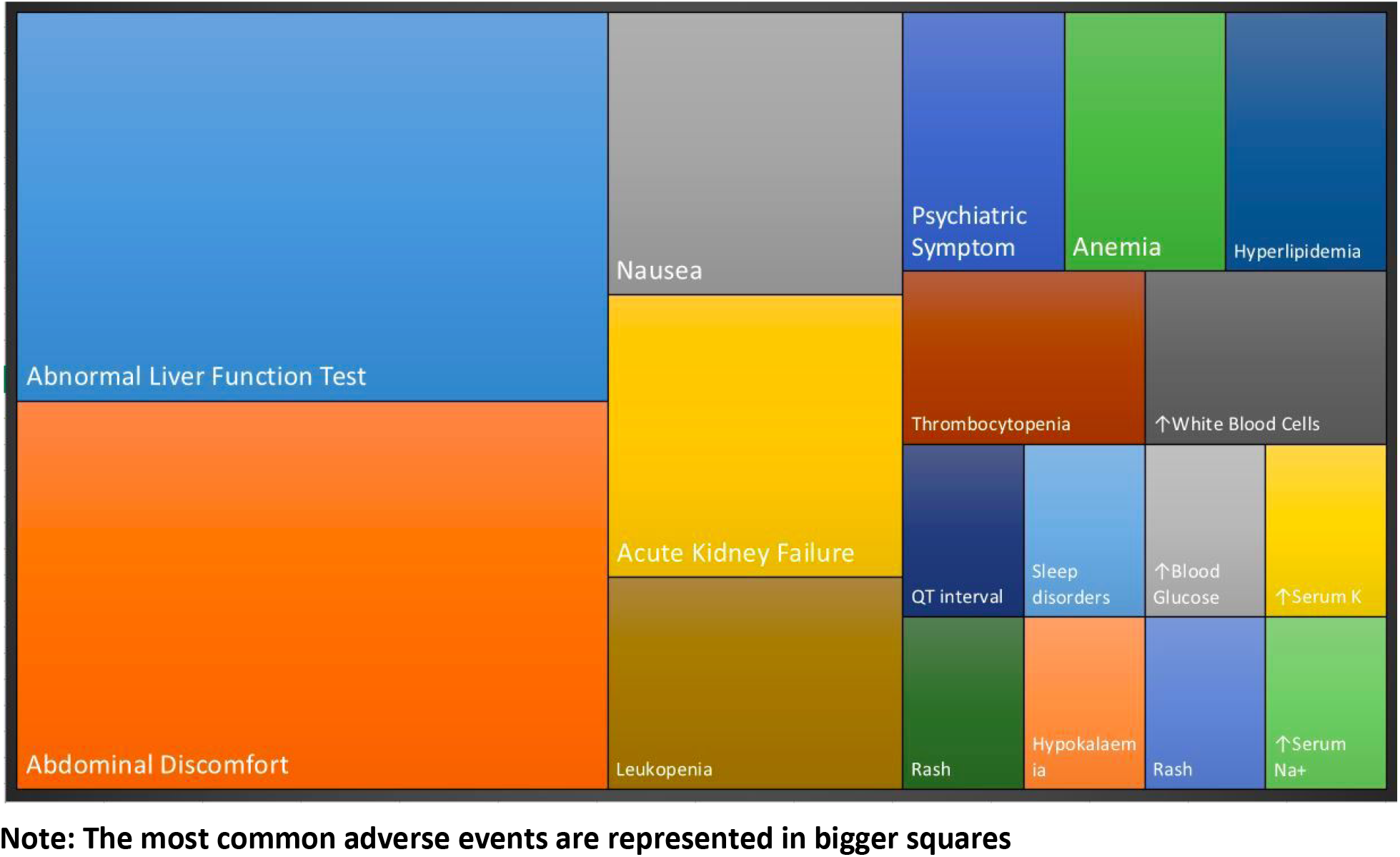
Most common adverse events within the studies

Cao et al reported AE of any grade in 48.4% of the patients in the LPV/r group and 49.5% in the standard care group (supplemental oxygen, noninvasive and invasive ventilation, antibiotic agents, vasopressor support, renal-replacement therapy, and extracorporeal membrane oxygenation (ECMO)). Gastrointestinal AEs were more common in the LPV/r group, but SAE were more common in the standard-care group (Cao et al., 2020). LPV/r treatment was stopped early in 13 patients (13.8%) because of AE.

Li et al reported that 35.3% had AEs in the LPV/r group when compared to 14.3% in the Umifenovir group (Li et al., 2020). One SAE occurred in the LPV/r group. Wang, et al reported 102 (66%) of 155 patients in the Remdesivir group and 50 (64%) of 78 in the control group. The Remdesivir group reported 28 (18%) and 20 (26%) in the control group (Wang et al., 2020). Beigel et al reported that SAE occurred in 114 patients (21.1%) in the Remdesivir group and 141 patients (27.0%) in the placebo group(Beigel et al., 2020). Chen et al reported a total of 37 (31.9%) of AEs in the Favipiravir group and 28 (23.3%) in the Umifenovir group (Chen et al., 2020).

### Clinical Improvement at day 7, 14, and 28

Cao et al reported that the percentage of patients with clinical improvement at day 7, 14 and 28 was higher in the LPV/r group than in the standard care group (day 7: 6% vs. 2%; day 14: 45.5% vs. 30.0%; day 28: 78.8% vs. 70.0%) (Cao et al., 2020). The study by Li et al showed no statistical differences of clinical improvement at day 7 nor at day 14 between the three groups (P > 0.05) (Li et al., 2020). Remdesivir use was not associated with a difference in time to clinical improvement (hazard ratio 1.23 [95% CI: 0.87–1.75]). The study showed a slightly higher improvement between the Remdesivir group compared to controls (day 7: 3% vs.3%; day 14: 27% vs. 23%; day 28: 65% vs.58%) (Wang et al., 2020).

### 28-day mortality

Mortality (28-day) was reported in only two studies (Cao et al., 2020; Wang et al., 2020). Cao et al showed some evidence that LPV/r reduces mortality at 28 days (19.2% vs. 25.0%; difference, −5.8 percentage points (95% CI, −17.3 to 5.7) (Cao et al., 2020). Wang et al reported similar 28-day mortality between the two groups, 14% for the Remdesivir group compared to 13% of the control groups; difference 1.1% (95% CI, –8.1 to 10.3). In patients with the use of Remdesivir within 10 days after symptom onset, 28-day mortality was not significantly different between the groups (Wang et al., 2020).

### Meta-analyses for RCTs

#### Antivirals vs controls

Four studies were included in the meta-analyses that compared antivirals vs placebo or standard of care(controls) of data (Beigel et al., 2020; Cao et al., 2020; Li et al., 2020; Wang et al., 2020). These encompassed 829 cases and 716 controls. There was no statistical difference in AE when comparing antivirals with controls (RR, 0.95 CI, 0.80 to 1.35, P= 0.52, I^2^=32%). However there was a significant difference with SAE (RR, 0.74, CI, 0.62 to 0.89, P= 0.002, I^2^=0.00]. Patients taking antivirals had 26% less risk of having a SEA compared to controls. This protective effect was not seen in the study that included a placebo as a control group.

There was no significant difference in the clinical improvement at day 7 and 28 among the cases treated with antivirals (RR, 1.09 CI, 0.55 to 2.15, P=0.80), (RR, 1.13 CI, 0.99 to 1.29, P=0.08] respectively. Clinical improvement at day 14 was observed among the cases treated with antivirals compared to the control group (RR, 1.24 CI, 1.00 to 1.53, p=0.05). Outcome related to 28-day mortality was not statistically significant among the cases and controls (RR, 0.87 CI, 0.57 to 1.33, P=0.52) (figure 2).

**Figure 2:**
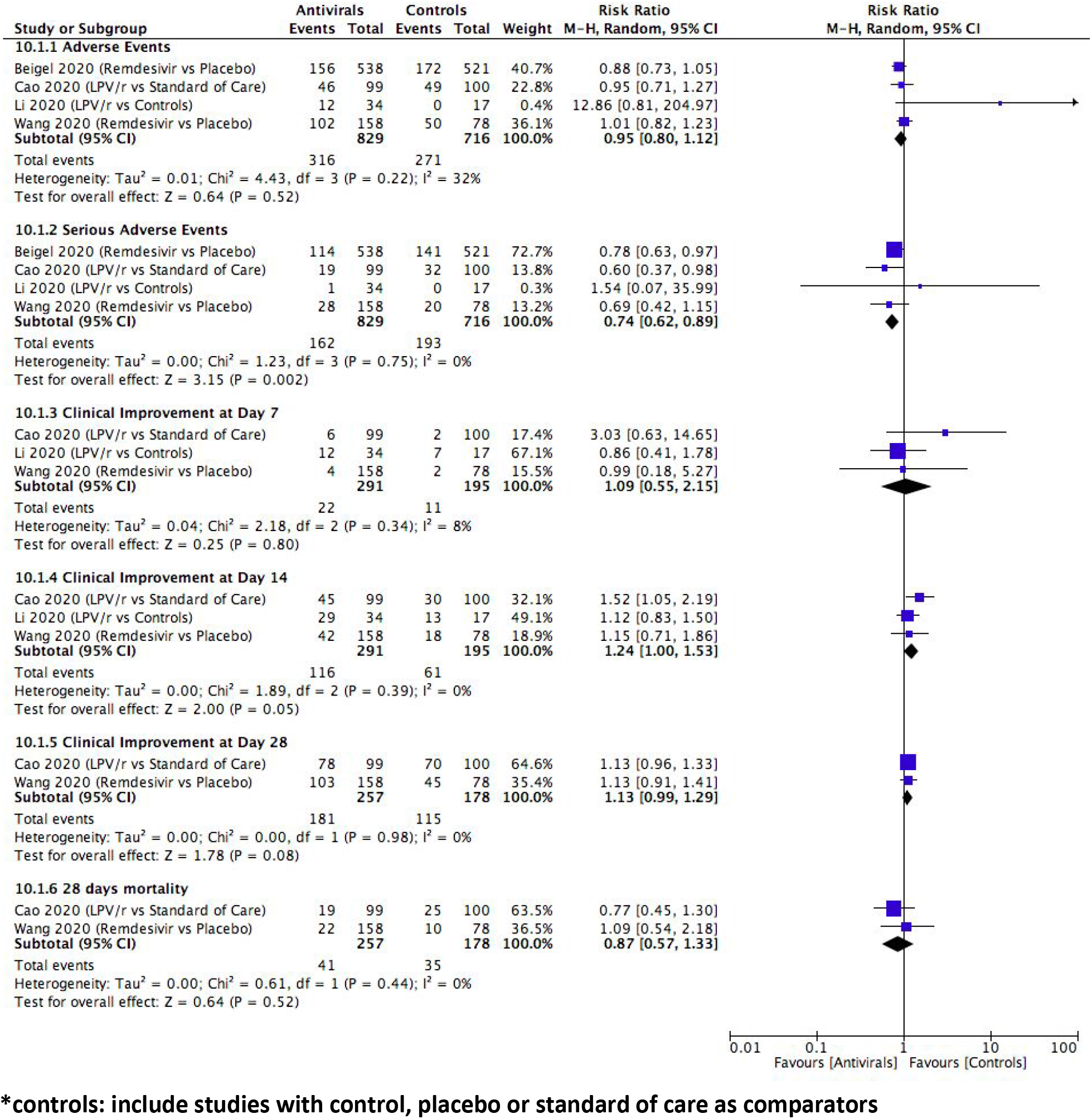
Forest Plot for RCTs showing the comparison of Antivirals vs Controls*

When stratifying the meta-analyses by specific antiviral there were no statistical differences between the antiviral and the control group. The heterogeneity and the confidence intervals were wide for the less SAE comparison, showing a lack of power.

### Meta-analysis for observational studies

Figures 5–7 (supplementary) show the meta-analyses in which there were two or more studies available. There was no statistical difference for nCOV negativity improvement at day 14 or mortality when comparing among the antivirals. Patients exposed to LPV/r had a chest improvement when compared to Umifenovir (RR 1.45, CI 1.04–2.04, p=0.03) (Deng et al., 2020; Lan et al., 2020; Liu et al., 2020; Zhu et al., 2020).

## DISCUSSION

The results of the meta-analyses do not show evidence of clinical improvement or reduced mortality for any antiviral agent in patients with severe and critical COVID-19. There is evidence from single studies that Remdesivir and LPV/r reduces the hospital length of stay. In addition, evidence derived from a meta-analysis of observational studies shows that patients exposed to LPV/r had chest X-Ray improvement when compared to Umifenovir. Concerning SAE it was observed that patients to which antivirals were administered had less SAE compared to patients without antivirals. The results of the meta-analyses should be interpreted with caution due to a lack of statistical power and a lack of high-quality RCTs.

The AEs reported with all antivirals were related to gastrointestinal symptoms and to abnormal liver function tests. SAE present in LPV/r and Remdesivir were acute kidney injury, acute respiratory distress syndrome, hemorrhage of the lower digestive tract, and septic shock. When interpreting these results one has to take into account that these outcomes might be associated with the symptoms of COVID-19 and its severity. Future studies need to determine if these outcomes are related to the disease or to the treatment.

Since all of the studies included in the meta-analyses had different types of limitations, it is important to focus on the systematic review and evaluate single studies with high quality. The study by Wang et al. was the only study with high quality. It was the first placebo-controlled, double-blinded RCT. Even though the study was conducted in ten hospitals, the study ended prematurely after including 237 patients due to slow recruitment, which meant the trial was underpowered and the outcome was inconclusive. The study showed that patients treated with Remdesivir within 10 days after the onset of clinical symptoms had a reduction of time on mechanical ventilation and a faster clinical improvement (2 days shorter) compared to standard therapy. In addition, the study by Beigel et al showed that Remdesivir shortened the time to recovery in hospitalized patients with COVID-19 (Beigel et al., 2020). Further clinical trial is warranted to clarify and strengthen the effect of Remdesivir on COVID-19 patients.

Concerning the results of LPV/r, it is important to note that the studies presented major limitations including the risk of bias and lack of power, therefore high quality is needed before a conclusion can be made. In vitro studies showed that LPC/r had antiviral activity against SARS-CoV (Choy et al., 2020; Li and De Clercq, 2020). In addition, the results of clinical studies reported that patients treated with LPV/r had a reduction in intubation rate and mortality with Lopinavir/Ritonavir in SARS patients.

Meta-analyses are used to increase the lack of statistical power when studies are small. Given that very few studies have been published we decided to include 2 pre-publications (non-peer-reviewed) (Lan et al., 2020; Liu et al., 2020). The advantage of including these studies is that it increases the size of the meta-analyses and reduces the risk of publication bias which occurs when positive results get published sooner than negative results. Sensitivity analyses were performed by removing these two studies, and the results did not change.

Given the sparse data that is available, it was not possible to conduct meta-analyses for each individual drug nor to study the heterogeneity. The difference between studies is that they included a range of treatment dosages, frequencies, and routes of administration. Some studies compared an antiviral with a placebo and other antivirals vs standard of treatment. Since a standard of treatment has not been determined, it is important that studies describe clearly what they are including as standard of treatment. In addition, there is evidence that different treatments might have different efficacies at different stages of illness, therefore studies should stratify by the severity of the disease instead of encompassing all patients (Wiersinga et al., 2020). It would be beneficial that after this pandemic, consortia developed an emergency template protocol that could be used in future infectious pandemics. By standardizing methodologies, definitions, and outcomes, the quality of meta-analyses would increase leading to faster treatment.

These first studies need to be taken as an initial exploration of a potential treatment for patients that are seriously ill and should only be taken as a first step to guide the treatment and research in the short term. As a medium-term plan, we are awaiting the results of large studies such as the WHO clinical trial SOLIDARITY (Kupferschmidt, 2020) and the Randomized Evaluation of COVID (RECOVERY) trial from the University of Oxford (Recovery Trial, n.d.) which will focus on the need for hospitalization or ventilation and will evaluate the survival (mortality). In the longer term, there is a need for large high-quality studies, which will carry more weight than meta-analyses that include lower quality studies. There is a need for large, well-controlled, randomized studies that besides evaluating safety and efficacy, can also answer questions. Some of the questions that need to be answered are: At what stage of the disease should antivirals be prescribed? At what dosages? Should more than one antiviral be prescribed? What patients (e.g. age, comorbidities, comedications) benefit the most? What treatments are safer for specific subpopulations (e.g., pregnant women, underlying COPD). There are many questions that still need to be answered, however, at this stage, systematic reviews and meta-analyses can be useful to lead the research and development of further studies.

## CONCLUSIONS

The results of this systematic review and meta-analyses have an overview of the first studies published and guide future studies on what should be considered when starting a clinical trial. There is evidence that Remdesivir and LPV/r reduces the hospital length of stay and that patients to which antivirals were administered had less SAE when compared to patients not prescribed with antivirals. Due to a lack of power and quality of the studies, it was not possible to make a definite conclusion, and therefore the optimal approach to antiviral treatment is still uncertain.

## Data Availability

All data is available in the supplementary document

## Disclaimer

SLL works at Novartis Pharmaceuticals. The views and opinions expressed in this article do not represent any official policy or position of Novartis and its affiliated companies.

## Disclaimer

Sandra Lopez Leon works at Novartis Pharmaceuticals. The views and opinions expressed in this article do not represent any official policy or position of Novartis and its affiliated companies.

